# Structure-based network analysis predicts mutations associated with inherited retinal disease

**DOI:** 10.1101/2023.07.05.23292247

**Authors:** Blake M. Hauser, Yuyang Luo, Anusha Nathan, Gaurav D. Gaiha, Demetrios Vavvas, Jason Comander, Eric A. Pierce, Emily M. Place, Kinga M. Bujakowska, Elizabeth J. Rossin

## Abstract

With continued advances in gene sequencing technologies comes the need to develop better tools to understand which mutations cause disease. Here we validate structure-based network analysis (SBNA)^1, 2^ in well-studied human proteins and report results of using SBNA to identify critical amino acids that may cause retinal disease if subject to missense mutation. We computed SBNA scores for genes with high-quality structural data, starting with validating the method using 4 well-studied human disease-associated proteins. We then analyzed 47 inherited retinal disease (IRD) genes. We compared SBNA scores to phenotype data from the ClinVar database and found a significant difference between benign and pathogenic mutations with respect to network score. Finally, we applied this approach to 65 patients at Massachusetts Eye and Ear (MEE) who were diagnosed with IRD but for whom no genetic cause was found. Multivariable logistic regression models built using SBNA scores for IRD-associated genes successfully predicted pathogenicity of novel mutations, allowing us to identify likely causative disease variants in 37 patients with IRD from our clinic. In conclusion, SBNA can be meaningfully applied to human proteins and may help predict mutations causative of IRD.

## Introduction

As genetic sequencing has become increasingly available and less costly, a growing number of patients with clinical presentations of suspected genetic origin are undergoing targeted or whole exome sequencing. Despite improved accessibility, the genetic basis of disease for a considerable proportion of these patients remains unclear following sequencing^3^. Inherited retinal diseases (IRD), whereby rod and cone photoreceptors degenerate, are a group of Mendelian disorders that represent an important causes of vision loss^4^. With the advancement in availability of genetic testing^5–8^, the lower cost of exome sequencing and the prospect of gene therapy for several diseases^8^, it is becoming a field with increasing promise and possibility. However, among patients with IRD, approximately 30% are not assigned a clear genetic basis despite classic retinal changes and decrease in visual and retinal function^9^. Though the field of IRD is one of the most advanced in terms of gene therapy trials, patients without a genetic diagnosis are not candidates for treatment. For these patients, additional tools are needed to better understand the phenotypic impact of identified genetic variants that are not among the group of known causal mutations (i.e. variants of uncertain significance -- VUS).

Numerous computational tools that aim to predict the phenotype of genetic variants have previously been described^10–12^, many of which were trained based on existing variant classifications or combine multiple metrics that use this type of training data^13–21^. These data can be limited by the sparsity of available annotations^22^, and previous studies suggest that some of these algorithms may have considerable false discovery rates^23, 24^. The clinical applicability of these approaches has been limited as a result^25^. Another class of methods uses sequence conservation to estimate the likelihood that a particular mutation will have a deleterious phenotypic effect^26, 27^. However, sequence conservation is an imperfect proxy for the functional importance of a particular amino acid position within a protein of interest; this has been previously demonstrated within the context of human immunodeficiency virus (HIV), which has a per-base mutation rate approximately 10^4^ times that of the human genome and thus serves as a model for an accelerated rate of genomic evolution^1, 28, 29^. Functional *in vitro* assays can be very helpful, such as a high content assay for the *RHO* gene to characterize genetic variants^30^. These approaches are crucial tools to test each variant, but on a large scale the setup is costly and not always feasible for the entire volume of patient-genotype combinations as they require the appropriate experimental setup, tissue-type and readout for the particular mutation of interest.

Structure-based network analysis (SBNA) leverages the application of network theory to protein structure data with the goal of quantifying local residue connectivity, bridging interactions, and ligand proximity in order to identify amino acid residues that are topologically important^1, 31^. Using x-ray crystallography data, it models proteins as networks of connected amino acids to quantitatively estimate the topological importance of each amino acid as it relates to others in the protein, protein complex or protein-ligand interaction. This approach is distinct from prior computational tools that use structural information because it is not reliant on pre-defined secondary structure elements; rather, it simply analyzes the crystallized tertiary structure of the folded protein as a network. Results from SBNA have been previously used to identify highly mutationally constrained CD8^+^ T cell epitopes within the context of both HIV and severe acute respiratory syndrome coronavirus 2 (SARS-CoV-2)^1, 2^. In the case of HIV, these epitopes were found to be preferentially targeted by individuals able to suppress HIV replication in the absence of antiretroviral therapy^1^. Within the context of SARS-CoV-2, highly networked regions in the virus were found to lack mutation in broadly circulating strains and were thus promising targets for mutation-resistant vaccinees potentially capable of conferring broad protection against sarbecovirus infection^2^.

Despite its promise in multiple viral families, SBNA has not been previously applied to human proteins which exist within a far more complex system of biological interactions. Such an application would potentially aid in a better understanding of which genetic variants discovered through sequencing are pathogenic. Since such a sizable number of patients with IRD harbor VUSs, we therefore sought to investigate whether this approach could generate meaningful results for IRD proteins of interest to estimate the phenotypic impact of missense mutations. After establishing that this approach could generate meaningful results in well-studied human proteins, we then applied SBNA to IRD genes in ClinVar, a large database of reported pathogenic and benign genetic variation. Finally, we addressed a cohort of 65 patients in our clinic at MEE who are affected by an IRD but for whom a genetic cause has yet to be discovered to investigate the use of SBNA in real-world clinical scenarios.

## Results

### Structure-based network analysis predicts mutations in human proteins associated with in vitro loss of function and pathogenic clinical phenotypes

Four well-studied human proteins – BRCA1, HRAS, PTEN, and ERK2 – were selected for analysis to evaluate whether the structure-based network analysis pipeline could be meaningfully applied to human proteins despite the increased complexity of humans compared to viruses. In addition to available high-quality full or partial structural data, *in vitro* saturation mutagenesis experiments were previously published for each of these proteins; we therefore extracted the functional consequence of all missense mutations^32–35^. We generated network scores for amino acid residues included in the available protein structures (**Fig. 1A**) and evaluated the correlation with the saturation mutagenesis functional scores (**Fig. 1B**). All four proteins showed a strong inverse correlation between network and functional scores, which was consistent with previous findings for viral proteins^1^.

**Figure 1.**
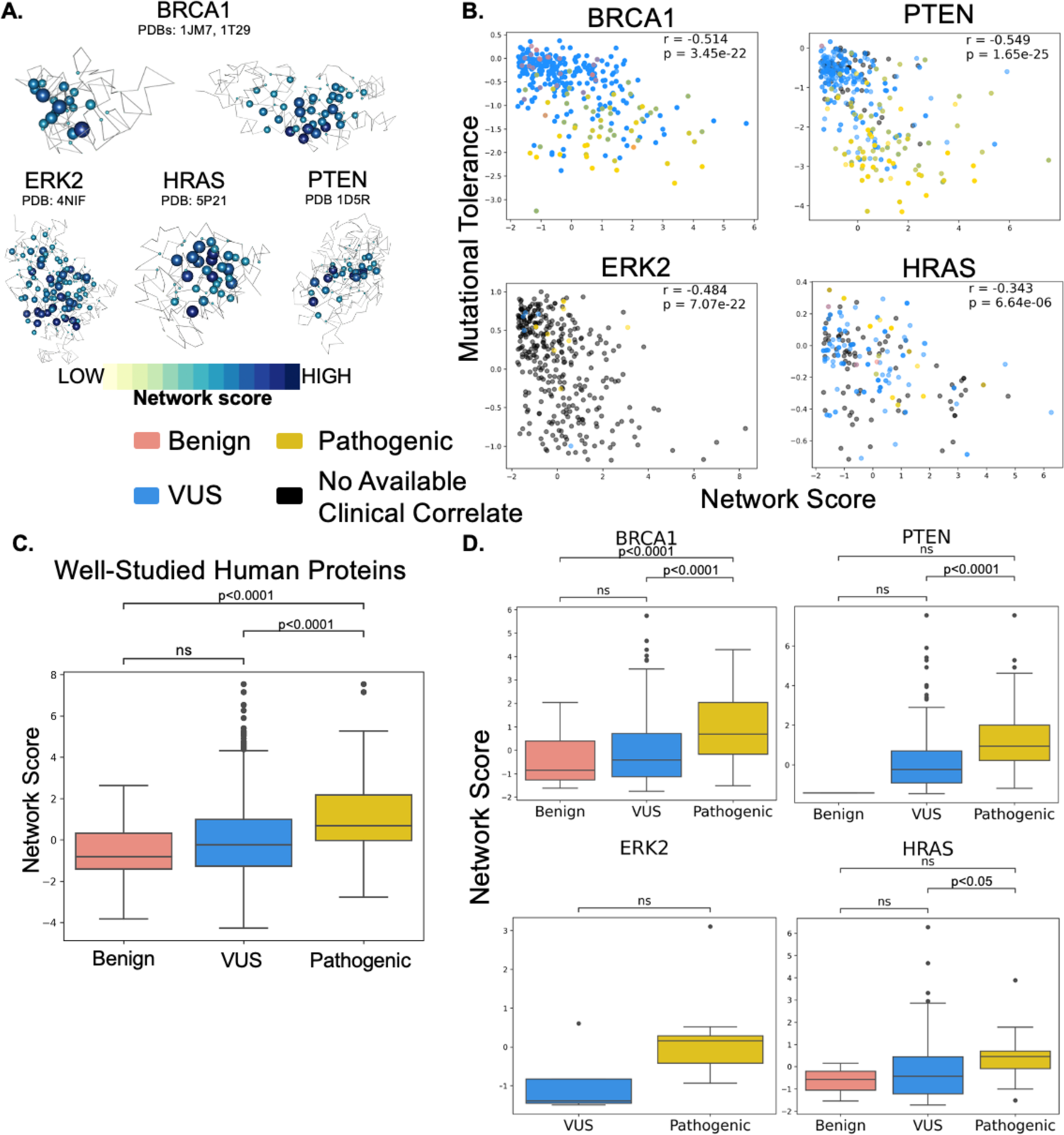
Structure-based network analysis highlights pathogenic variants in well-studied human proteins. **(A)** Structural representations show network scores at each residue. Sphere radius corresponds to network score magnitude at a particular position. **(B)** Comparison between functional data from saturation mutagenesis experiments and network scores, with Spearman correlation coefficients and p-values displayed for each plot. Points are colored based on available clinical phenotype data. **(C)** Pooled comparison between network scores for variants with available clinical phenotype data for all four well-studied human proteins. **(D)** Individual comparisons between network scores for variants with available clinical phenotype data for all four well-studied human proteins.

We next compared network scores to pathogenicity categorizations derived from human data using the ClinVar and gnomAD genetic databases for these same four proteins (**Fig. 1C**). Missense variants in all subsequent analyses were categorized with respect to human clinical data in line with the American College of Medical Genetics and Genomics (ACMG)^25^ as benign (encompassing ‘benign’ or ‘likely benign’ within ClinVar), VUS or pathogenic (encompassing ‘pathogenic’ or ‘likely pathogenic’ within ClinVar). We restricted our analysis to ClinVar missense mutations with at least two-star level evidence, and gnomAD was used to identify relatively benign missense mutations (variants with ≥1% allele frequency which are unlikely to be the driver in an IRD). Across all four proteins, network scores assigned to pathogenic variants were significantly greater than those assigned to benign variants (median network scores for benign missense mutations −0.936 and pathogenic mutations 0.866, P=1.836e-5, **Fig. 1C**). Scores assigned to VUSs fell in between those assigned to benign and pathogenic variants. Looking more closely at network scores corresponding to individual proteins, the trend of pathogenic variants being assigned greater network scores than benign variants held, though this was not statistically significant in smaller sample sizes (**Fig. 1D**). Overall, network scores correlate with available clinical phenotype data for the four well-studied human proteins (Spearman correlation coefficient 0.228, p=2.116e-23), suggesting that structure-based network analysis can be meaningfully applied to human proteins despite the complexity of the biological systems within which they exist.

### Structure-based network analysis predicts mutations in inherited retinal disease genes associated with pathogenic clinical phenotypes

Having established that structure-based network analysis could be meaningfully applied to 4 canonical well-studied human proteins with saturation mutagenesis data, we analyzed the relationship between network scores and pathogenicity designations from high-quality ClinVar variants and benign gnomAD variants (≥1% allele frequency) for 47 human genes associated with IRD. We found that pathogenic mutations were assigned significantly greater network scores compared to both benign mutations and VUSs (median benign −0.841, median VUS - 0.188, median pathogenic 0.947 p=3.140e-29 for benign vs. pathogenic, p=1.753e-60 for VUS vs. pathogenic; **Fig. 2A**), which mirrors the pattern previously observed in the well-studied canonical human proteins. The magnitude of these differences was robust in the setting of differing levels of available clinical data across genes up to a point: the results were consistent if restricting to genes with fewer than 100 high-quality variants, and the distinction between benign and pathogenic was detectable down to 40 high-quality entries per gene (**Fig. 2B-D**).

**Figure 2.**
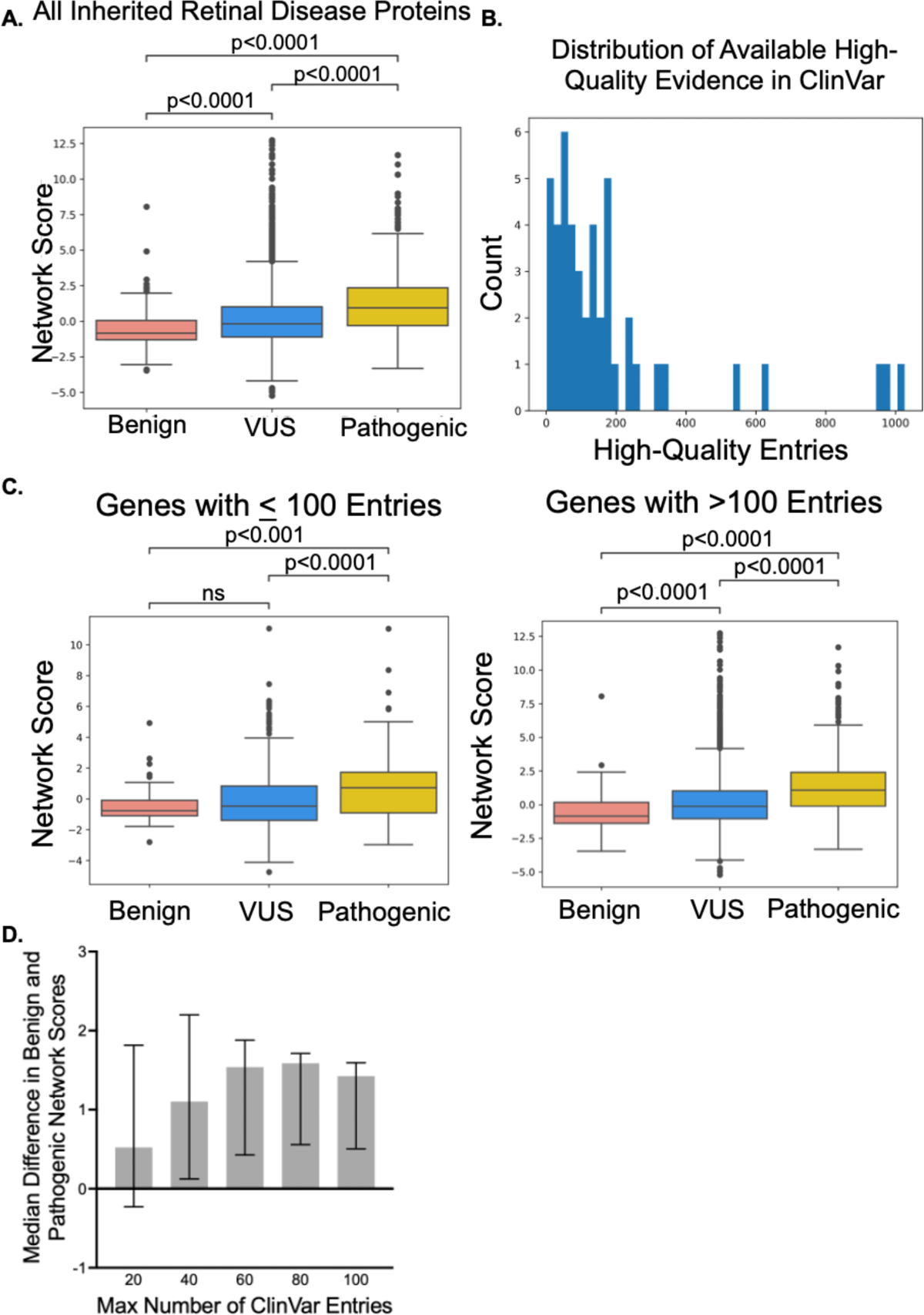
Structure-based network analysis highlights pathogenic variants in inherited retinal disease proteins. **(A)** Pooled comparison between network scores for variants with available clinical phenotype data for all 47 inherited retinal disease proteins. **(B)** Distribution of available high-quality evidence in ClinVar across all 47 inherited retinal disease proteins. **(C)** Comparison between network scores and clinical phenotypes grouped by level of available high-quality evidence in ClinVar for each inherited retinal disease protein. **(D)** The statistical significance of the difference in network scores between benign and pathogenic variants is lost between 20 and 40 high-quality ClinVar evidence entries.

Network score comparisons were also considered at the level of individual genes, as aggregate metrics provide limited clinical insights. For those genes with at least one benign variant and one pathogenic variant, 28/30 contain pathogenic variants with higher median network scores compared to benign variants (**Fig. S1**). However, the ability to assess statistical significance is limited on a per-gene basis due to fewer high-evidence mutations.

### Incorporating structure-based network analysis scores into multivariable logistic regression models successfully predicts variant pathogenicity

To facilitate the prediction of pathogenicity using network scores, we constructed a multivariable logistic regression model that incorporates not only the importance of a particular position within a protein (the SBNA score) but also the degree of chemical dissimilarity between the reference and mutant amino acid at that position. To capture the latter effect, we used the BLOSUM62 matrix as a covariate that accounts for the chemical similarity of a given amino acid change^36^. Models with network scores alone, BLOSUM62 scores alone, and both network and BLOSUM62 scores were tested for each of the three datasets previously considered in pathogenicity comparisons.

For each logistic regression model, we tested 500 iterations of a 70%/30% train-test split and calculated receiver operating characteristic (ROC) curve statistics for high-quality ClinVar variants and benign gnomAD variants. Based on mean area under the curve (AUC), models incorporating network scores outperformed BLOSUM62 alone. The logistic regression model incorporating both network scores and BLOSUM62 scores had the best performance across the inherited retinal disease genes (AUC 0.835, **Fig. 3A**). Based on these results, we determined that the regression with both network and BLOSUM62 scores was most likely to provide clinically relevant pathogenicity predictions. A leave-one-out logistic regression training approach displayed a similar pattern (**Fig. S2**).

**Figure 3.**
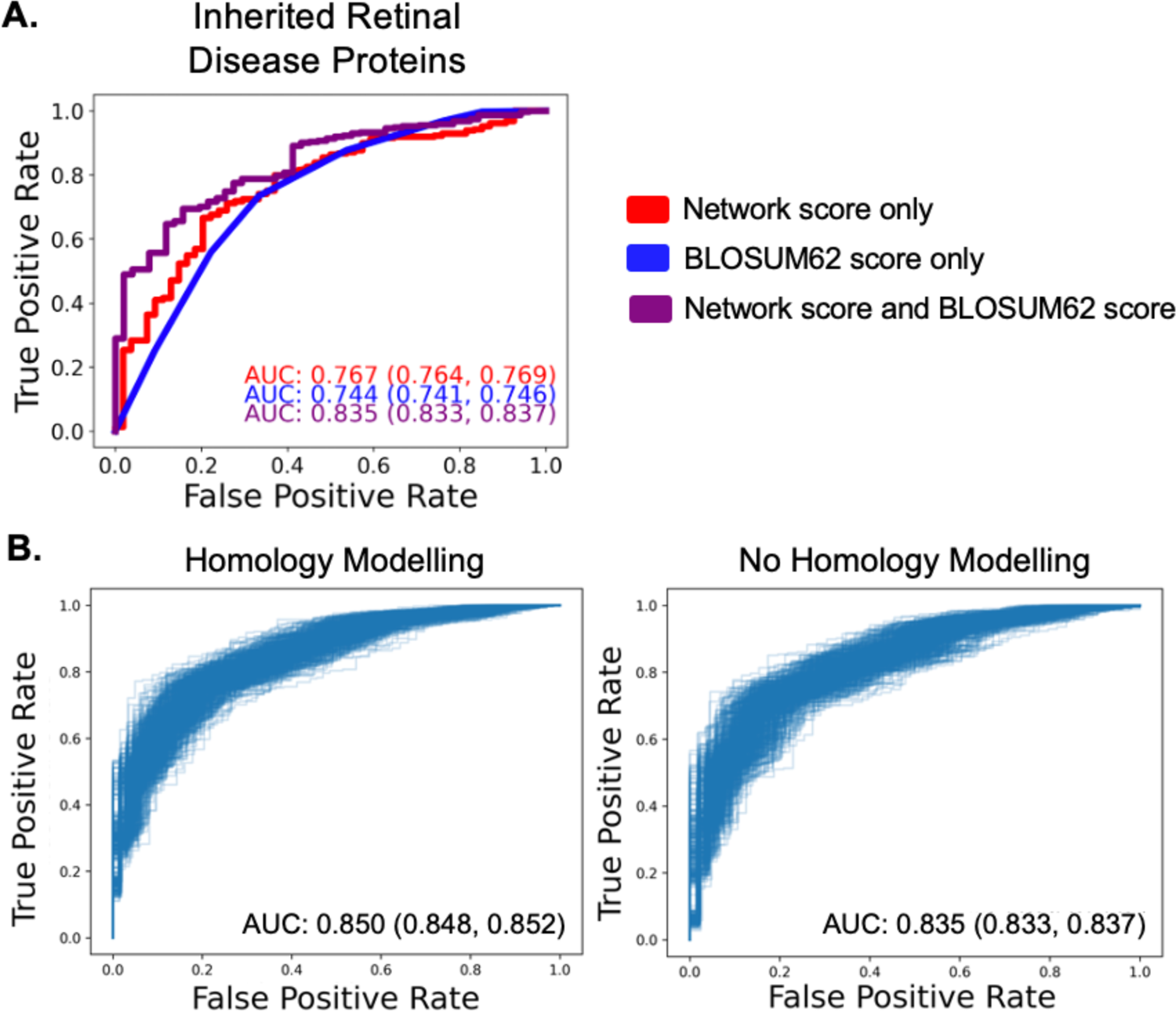
Logistic regression-based modelling using SBNA and BLOSUM62 is superior to univariate models. **(A)** Application of univariate and multivariable logistic regression to the inherited retinal disease protein datasets. All regressions were performed using a 70%/30% train/test data split with 500 iterations, and a representative ROC curve with AUC closest to the mean is shown for each regression model. AUC values are displayed as the mean followed by a 95% confidence interval. **(B)** Comparison of homology modelling versus no homology modelling over 500 iterations of a 70%/30% train/test data split for the inherited retinal disease protein dataset. ROC curves for each iteration are shown.

Polyphen2 is currently the most widely used computational prediction tool for variant pathogenicity^37^. Because PolyPhen2 is used by the ACMG to assign the pathogenicity designations that then are used in databases such as ClinVar and HumDiv, comparing the performance of SBNA to PolyPhen2 is inherently biased. We therefore decided to test whether SBNA predictions aligned with Polyphen2. Pathogenicity probability estimates from the 70%/30% train-test split regression including both SBNA and BLOSUM62 scores were compared to results generated using PolyPhen2 trained on two different datasets, HumDiv and HumVar^14, 37^. The SBNA regression pathogenicity probability estimates assigned to both benign and pathogenic variants as determined by ClinVar and gnomAD correlated with the PolyPhen2 probability estimates, but the PolyPhen2 results clustered more strongly at 0 and 1 (**Fig. S3A**). Despite this, the HumDiv-trained PolyPhen2 results, the HumVar-trained PolyPhen2 results, and the structure-based network analysis regression results showed a significant difference between the pathogenicity probabilities assigned to known pathogenic variants as compared to known benign variants (p=9.066e-64, p=2.747e-58, p=4.781e-44, respectively; **Fig. S3B**). The Spearman correlation between SBNA probabilities and PolyPhen2 probabilities was 0.445 for the HumDiv-trained PolyPhen2 results (p=5.13e-57) and 0.476 for the HumVar-trained PolyPhen2 results (p=4.11e-66).

### Homology modeling

As crystal structures are snapshots in time, we tested whether homology modeling, an *in silico* approach that allows proteins conformational flexibility, improves the performance of the SBNA pipeline. In addition, homology modeling has the added benefit of predicting human structure from non-human protein data. We implemented the well-documented tool Modeller^38–41^, and network scores were averaged across 50 homology models (**Fig. S4A**). Homology modelling yielded consistent improvement in the correlation between *in vitro* functional data across 9 bacterial and viral proteins that were previously used to benchmark the SBNA algorithm^1^ (**Fig. S4B**). We then applied this model to the 47 IRD genes. There was a statistically significant improvement in AUC with homology modelling as compared to without homology modeling (non-homology modeled AUC = 0.835 (95% confidence interval: 0.833, 0.837), homology modeled AUC = 0.850 (95% confidence interval: 0.848, 0.852), p < 0.0001, **Fig. 3B**). However, this difference is relatively small in magnitude and is unlikely to be clinically significant. Furthermore, this improvement was not consistently observed across individual proteins (**Fig. S5**). We therefore used the regression model fitted to the data without homology modeling for downstream analysis.

### Structure-based multivariable logistic regression model solves patients with unclear genetic basis for clinical disease

A significant percentage of patients with clinical presentations consistent with inherited retinal diseases (IRD) lack an identified genetic basis for their phenotype, and this pattern holds within the set of patients who receive care at Massachusetts Eye and Ear (MEE). We evaluated 455 patients who were diagnosed with an IRD at MEE based on visual acuity, visual field testing, clinical exam, fundus autofluorescence imaging, optical coherence tomography and electroretinogram and underwent targeted or whole exome-sequencing but lacked the canonical genetic mutation pattern to explain clinical findings. Mitochondrial causes of IRD were excluded. Missense variants of interest were defined based on allele frequency and residence in one of 250 known IRD genes. There were 3,727 identified variants in total (2,948 unique variants) residing in 51 genes and these were categorized as either pathogenic, “likely pathogenic” (LP), VUS or benign based on known mutational consequence in the literature using ACMG criteria^42^ and ClinVar designations^22^ (**Fig. S6**). Variants were further categorized in the context of individual patients as “likely solving” if they were pathogenic or LP, the clinical presentation was consistent with the known consequence of the affected gene and the zygosity was consistent with known modes of inheritance. Of the 455 patients reviewed, before applying SBNA, 355 were found to have variants that were “likely solving” while 65 patients harbored one or more VUSs that prohibited a molecular diagnosis. The remaining 35 patients had non-missense variants as well as missense variants or variants in a protein or region of a protein without available structural data. Therefore, SBNA could not be applied comprehensively to these patients.

Using a multivariable logistic regression model incorporating both network and BLOSUM62 scores that was trained on ClinVar data for the 47 IRD genes, we generated pathogenicity predictions for the 2,948 genetic variants identified in all 455 patients (**Fig. 4A**). Substitutions were then considered based on likelihood of pathogenicity determined using the fitted SBNA multivariable logistic regression model, and these pathogenicity probabilities were considered in the context of the identified genetic variants in known IRD genes for each patient with a clinical presentation consistent with IRD. Variants were matched with any phenotypic data available in ClinVar to roughly benchmark the quality of the pathogenicity likelihood estimates.

**Figure 4.**
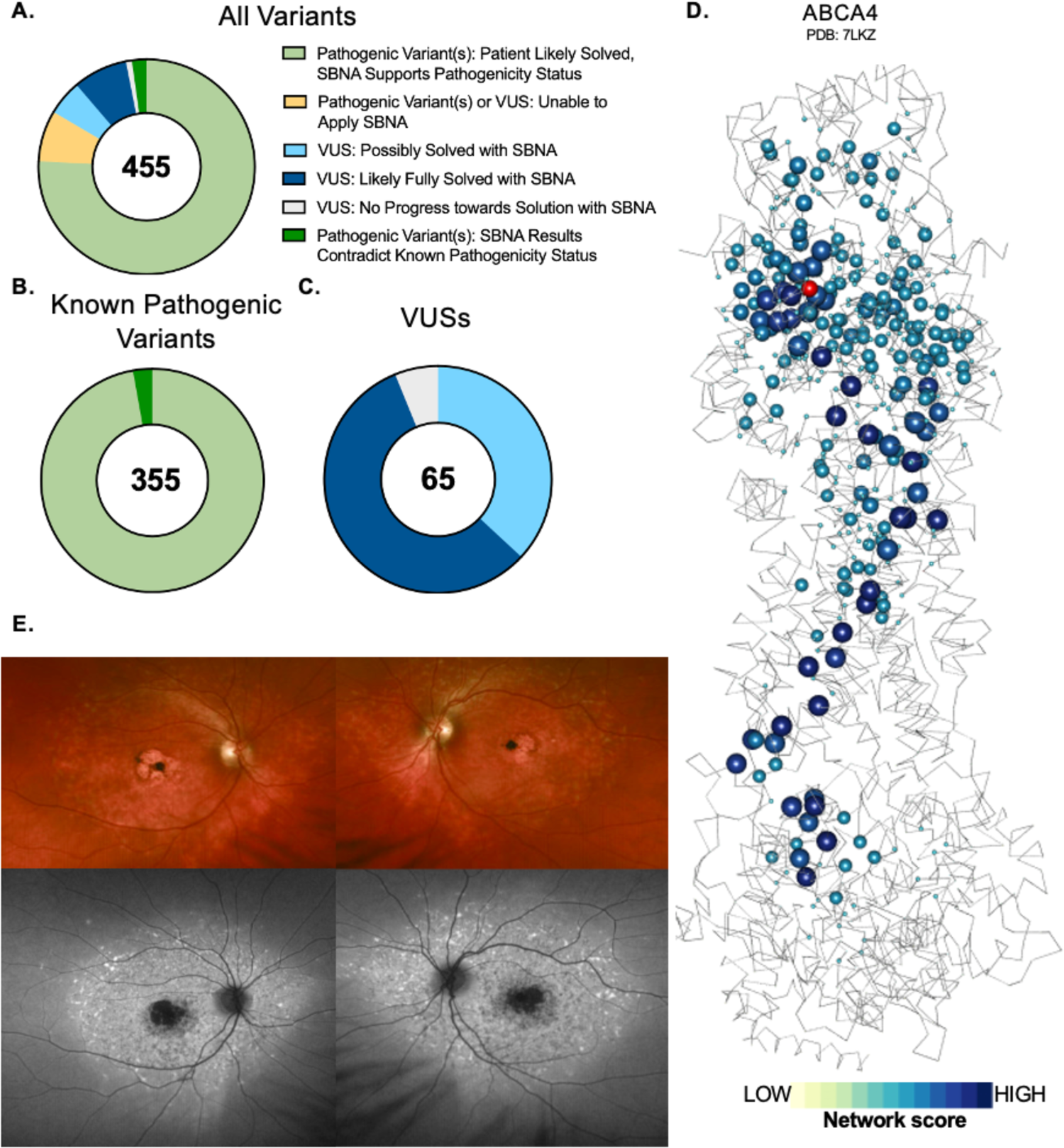
SBNA helps identify pathogenic variants in patients with inherited retinal disease. **(A)** Categorization of results from application of structure-based network analysis (SBNA) regression to a dataset of possibly solving patient variants. Results were further subdivided into those from patients with known putative genetic causes of disease **(B)** and those from patients with only VUSs in known inherited retinal disease-associated genes **(C)**. **(D)** Representation of network scores for a sample structure with putative solving genetic mutations. Sphere radius corresponds to network score magnitude at a particular position. A patient with clinical evidence of ABCA4 disease **(D)** but with no complete genetic explanation was fully solved using SBNA which highlighted two mutations (Pro1380Leu and Arg1097Ser) that score highly in the ABCA4 protein structure **(E)**. Arg1097Ser was a VUS and is indicated in red within the structure.

The distribution of pathogenicity probabilities assigned to known pathogenic and benign variants is shown in **Fig. S7A-B**. Similar to the ClinVar analysis of IRD genes, there were observable differences between the SBNA pathogenicity probabilities assigned to known pathogenic variants as compared to benign variants and VUS/variants without any available clinical data (benign median 0.352, VUS median 0.645, pathogenic median 0.958; benign vs. pathogenic p=2.528e-26). Rankings were performed using a multivariable logistic regression model fitted to the IRD-associated gene data with and without homology modelling, and the results were found to be highly coordinated (**Fig. S7**).

For the 355 patients who harbored known pathogenic variants sufficient to cause disease, the SBNA pathogenicity probability estimates were concordant with these pathogenicity categorizations in 97.2% of cases (**Fig. 4B**). For the 65 patients with VUSs as categorized by ACMG standards^42^ and/or ClinVar who were not already solved, the SBNA probability estimates offered support for a genetic cause of disease for 37 patients (22 unique variants, **Fig. 4C-E**).

Modes of inheritance included autosomal recessive (in combination with a known pathogenic variant or a second VUS with a high estimated probability of pathogenicity; n = 15), autosomal dominant (n = 2), and X-linked recessive (n = 5). For example, a patient with autosomal recessive *ABCA4*-related disease (typically a recessive inheritance pattern) was found to have variants Pro1380Leu (known pathogenic) and Arg1097Ser (VUS). Arg1097Ser scored highly within the multivariable logistic regression model (SBNA score 3.672, BLOSUM62 score −1, pathogenicity probability 0.965), suggesting it is likely pathogenic and thus completing the genetic solution for this patient. Similarly, the VUS Cys302Tyr in *RPGR* was found in a male hemizygous patient with phenotypic findings consistent with X-linked IRD and also scored highly within the multivariable logistic regression model (SBNA score 2.154, BLOSUM62 score −2, pathogenicity probability 0.956) (**Fig. S8**). The structure-based network analysis regression results may also have contributed towards identifying a possible genetic cause for an additional 24 patients. In these cases, there may have only been one identified heterozygous VUS, or there were multiple VUS identified but an SBNA score could only be generated for one of them due to the limitations of the available structural data. Finally, for the remainder of patients, SBNA was not possible due to lack of crystal structure data for these proteins or due to the presence of non-missense mutations.

## Discussion

In this study, we demonstrated that applying SBNA to human proteins yields results that correlate strongly with existing clinical phenotype data. We leveraged those scores, in combination with BLOSUM62 substitution scores, to generate estimates of the likelihood that a particular missense mutation was pathogenic. Using these estimates, we were able to nominate putative genetic solutions for 37 patients with clinical evidence of IRD. These results suggest that SBNA may be able to provide similarly meaningful insights for patients with an unclear genetic basis for their clinical symptoms, particularly for patients with inherited retinal diseases.

Numerous computational tools for prediction of variant pathogenicity have been developed^17, 43–54^. Virtually all of these tools use sequence conservation and protein secondary structure or domain information. These two categories of features work well, especially in conjunction with one another, because they harness both a nonspecific but sensitive measure (sequence conservation) and a more specific measure of structural constraint. However, SBNA is distinct in that it captures the structural topology of individual amino acids in the context of the protein as a whole and does not rely on pre-existing annotations. With so many methods available now, it is important to note that none of these tools will independently capture the phenotypic consequence of all coding mutations in human disease. Rather, the most successful approach likely will be to incorporate information from several tools when analyzing patient data.

Strengths of this approach include the lack of dependence on pre-existing clinical phenotype labels to generate network scores, which limits susceptibility to bias^22^. However, this approach is limited by the availability of high-quality structural data, a requirement for SBNA^1^. Numerous proteins, including the majority that correspond to known genetic variants associated with IRD, lack available x-ray crystallography or cryogenic electron microscopy structures altogether. In some cases, structures are available but are not of sufficient resolution to facilitate downstream network analysis. Furthermore, this approach will only be applicable to mutations that result in a negative structural change. Other types of mutations – such as splice site mutations – will not be captured with this approach.

Software that leverages artificial intelligence to predict protein structure, such as AlphaFold or RoseTTAFold, could potentially help to compensate for the lack of available structural data^55, 56^. However, these approaches remain highly limited in their ability to capture biological interactions between proteins^57^, and in our experience the performance of *de novo* structure predictions is poor in cases where a homologous structure does not already exist. While even high-quality structural data faces practical limitations in terms of the extent to which it can accurately represent the full spectrum of biological interactions, forgoing this valuable information entirely limits the utility of these structures for downstream applications such as the one described here. We therefore restricted our analysis to structural data available in the Protein Data Bank, but this in turn limited the number of proteins we were able to consider for this analysis.

In conclusion, SBNA provides an alternative approach to estimating the likely extent of phenotypic impact of protein variants that relies on modelling intramolecular and intermolecular interactions. We demonstrated that this technique could be meaningfully applied to human proteins and showcased the use of SBNA in IRD patients who lack a clear genetic diagnosis. Looking ahead, these types of insights could contribute to the design of novel gene therapies targeted at implicated genetic variants^58–60^.

## Methods

### Structural Data

All structural data was downloaded from the Protein Data Bank^31^. Individual accession numbers for the well-studied human proteins and inherited retinal disease proteins are listed in **Table S1**. For the non-human benchmark proteins, the same files were used as previously described^1^. In cases where multiple human structures were available, the highest oligomeric state of the protein with resolution of ∼3 angstroms or better was used. In cases where no human structure was available, the structure of one or more homologs was used. If multiple chains were present within the PDB file due to crystal packing rather than true oligomerization, only one of these chains was used for SBNA. Solvent and water molecules were removed from all PDB files prior to SBNA, but ligands and protein binding partners were included in the analysis. Only he protein of interest was designated to have a network score calculated by SBNA.

### Structure-based Network Analysis

Structure-based network analysis was used to calculate network scores as previously described^1, 2^. The details of this method have been described previously. As before, in cases where multiple conformations of a structure were used, network scores were averaged for each amino acid position. Where indicated, homology modelling was implemented using the previously described Modeller package^38–41^. In cases where no human structure was available, the structures of one or more non-human homologs as listed in **Table S1** were inputted into Modeller. When homology modelling was not used, only one of these homologous structures was considered.

### Data Analysis and Visualization

Data analysis was performed using Python (version 3.8.2), with visualizations generated using the “matplotlib” package. Logistic regressions were performed using the “glm” package in R (version 4.0.4). Intercepts were set to zero for all logistic regression models. Network score visualizations were generated in R (version 4.0.4) using the “rgl” package to implement OpenGL. The backbone centroid (nitrogen, alpha carbon, and carbon) positions were plotted along x, y, and z axes, and nodes were colored and given sphere radii corresponding to network scores. The protein structure backbone was then plotted, connecting the alpha carbons. Plotted structures were manually rotated, and two-dimensional views of interest were downloaded for inclusion.

### Phenotype Data

Clinical phenotype data was downloaded from ClinVar. Clinical evidence at with a 2-gold star level designation (meaning that “two or more submitters with <u>assertion criteria</u> and evidence (or a public contact) provided the same interpretation”) or better was included in the published analyses^22^. Pathogenicity designations from ClinVar were binned into Benign (“Benign”, “Benign/Likely benign”, “Likely benign”), VUS (“Uncertain significance”, “not provided”, “Conflicting interpretations of pathogenicity”), and Pathogenic (“Pathogenic”, “Pathogenic/Likely pathogenic”, “Likely pathogenic”) categories for analysis. Additionally, allelic variation data from gnomAD^61^ was considered for each gene. Loci with at least 250 available allelic variants in gnomAD were considered as benign variants. Results from ClinVar and gnomAD for each gene were considered in combination for these analyses.

#### Functional Data

Functional data for the four well-studied human proteins – BRCA1^32^, ERK2^35^, PTEN^33^, and HRAS^34^ – that had been previously published was used for this analysis. In cases where functional scores were assigned to multiple amino acid mutations at the same position (e.g., if different functional scores were calculated for Ala101Pro and Ala101Gln), the arithmetic mean of all functional scores at that position was used.

For the non-human benchmark proteins used to analyze the impact of homology modelling, functional data was also obtained from previously published data^62–74^. These data were processed for comparison with network scores as previously described^1^.

#### Patient Data

Patient data was gathered from among those presenting to the Inherited Retinal Disorders Service at Massachusetts Eye and Ear. Appropriate consent was obtained from all included patients, and ethical approval was granted by the Mass General Brigham/Massachusetts Eye and Ear Institutional Review Board.

#### Statistical Analysis

Statistical analysis, including generation of ROC curves, was performed using the “scipy.stats” package in Python as well as GraphPad Prism (version 9). Comparisons between three or more categories were made using the non-parametric Kruskal-Wallis test with Dunn’s post hoc analysis corrected for multiple comparisons with a Bonferroni correction. Comparisons between two categories were performed using the non-parametric Mann-Whitney U test. Correlation between two datasets was calculated using non-parametric Spearman correlation coefficients. Spearman correlation coefficients between network scores and ClinVar pathogenicity designations was calculated by assigning 0 to benign variants, 1 to VUS, and 2 to pathogenic variants.

## Data Availability

All data produced in the present study are available upon reasonable request to the authors.

## Notes

**Funding:** B.M.H. is supported by award number T32GM144273 from the National Institute of General Medical Sciences and award number F30AI160908 from the National Institute of Allergy and Infectious Diseases. E.J.R. is supported by award number K12EY016335 from the National Eye Institute and the Massachusetts Lions Eye Research Fund. The funding organizations had no role in design or conduct of this research.

### Competing Interest Statement

E.J.R. and G.D.G. have filed patent application PCT/US2021/028245.

### Funding Statement

B.M.H. is supported by award number T32GM144273 from the National Institute of General Medical Sciences and award number F30AI160908 from the National Institute of Allergy and Infectious Diseases. E.J.R. is supported by award number K12EY016335 from the National Eye Institute and the Massachusetts Lions Eye Research Fund. The funding organizations had no role in design or conduct of this research.

### Author Declarations

The Institutional Review Board of Mass General Brigham gave ethical approval for this work.

