## Supplement for "Structure-based network analysis predicts mutations associated with inherited retinal disease"

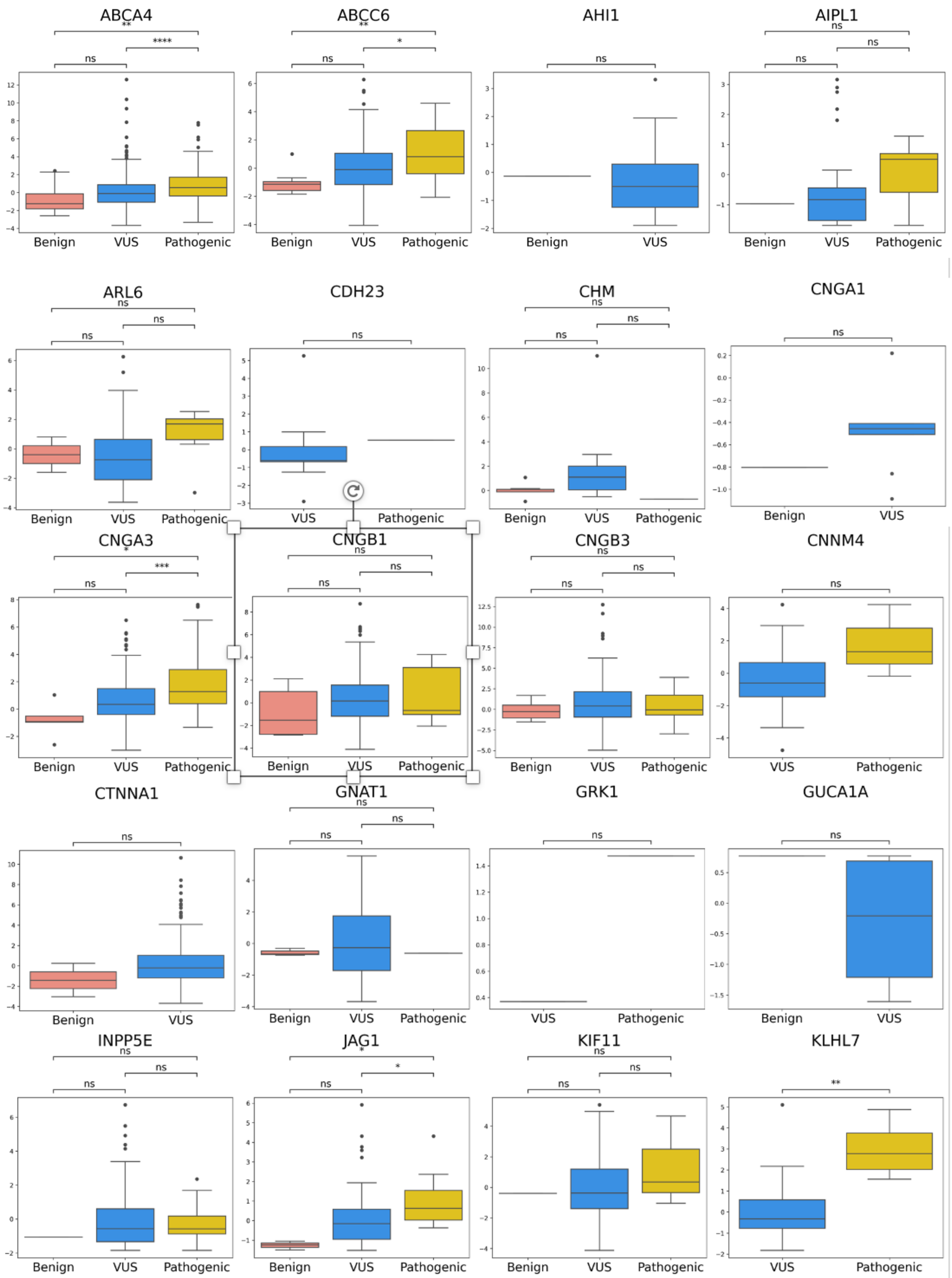
ABCA4
**
****
ns
Benign
VUS
Pathogenic
ABCC6
**
*
ns
Benign
VUS
Pathogenic
AHI1
ns
Benign
VUS
AIPL1
ns
ns
ns
Benign
VUS
Pathogenic
ARL6
ns
ns
ns
Benign
VUS
Pathogenic
CDH23
ns
VUS
Pathogenic
CHM
ns
ns
ns
Benign
VUS
Pathogenic
CNGA1
ns
Benign
VUS
CNGA3
*
***
ns
Benign
VUS
Pathogenic
CNGB1
ns
ns
ns
Benign
VUS
Pathogenic
CNGB3
ns
ns
ns
Benign
VUS
Pathogenic
CNNM4
ns
VUS
Pathogenic
CTNNA1
ns
Benign
VUS
GNAT1
ns
ns
ns
Benign
VUS
Pathogenic
GRK1
ns
VUS
Pathogenic
GUCA1A
ns
Benign
VUS
INPP5E
ns
ns
ns
Benign
VUS
Pathogenic
JAG1
*
*
ns
Benign
VUS
Pathogenic
KIF11
ns
ns
ns
Benign
VUS
Pathogenic
KLHL7
**
VUS
Pathogenic

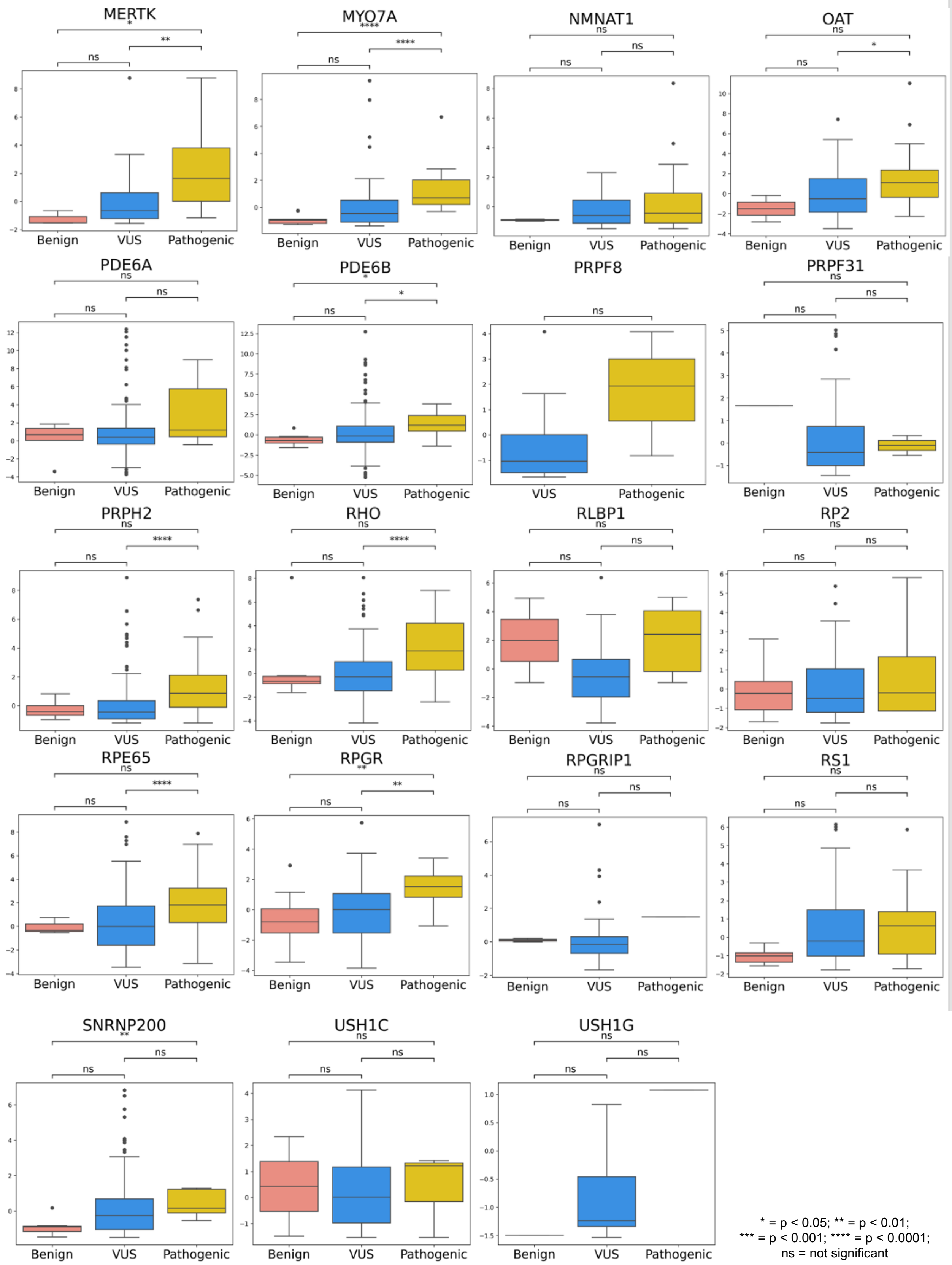
MERTK
*
**
ns
Benign
VUS
Pathogenic
MYO7A
****
****
ns
Benign
VUS
Pathogenic
NMNAT1
ns
ns
ns
Benign
VUS
Pathogenic
OAT
ns
*
ns
Benign
VUS
Pathogenic
PDE6A
ns
ns
ns
Benign
VUS
Pathogenic
PDE6B
*
*
ns
Benign
VUS
Pathogenic
PRPF8
ns
VUS
Pathogenic
PRPF31
ns
ns
ns
Benign
VUS
Pathogenic
PRPH2
ns
****
ns
Benign
VUS
Pathogenic
RHO
ns
****
ns
Benign
VUS
Pathogenic
RLBP1
ns
ns
ns
Benign
VUS
Pathogenic
RP2
ns
ns
ns
Benign
VUS
Pathogenic
RPE65
ns
****
ns
Benign
VUS
Pathogenic
RPGR
**
**
ns
Benign
VUS
Pathogenic
RPGRIP1
ns
ns
ns
Benign
VUS
Pathogenic
RS1
ns
ns
ns
Benign
VUS
Pathogenic
SNRNP200
**
ns
ns
Benign
VUS
Pathogenic
USH1C
ns
ns
ns
Benign
VUS
Pathogenic
USH1G
ns
ns
ns
Benign
VUS
Pathogenic
* = p < 0.05; ** = p < 0.01;
*** = p < 0.001; **** = p < 0.0001;
ns = not significant

**Figure S1. Structure-based network analysis highlights pathogenic variants in individual inherited retinal disease proteins.** Select individual comparisons between network scores for variants with available clinical phenotype data for inherited retinal disease proteins.

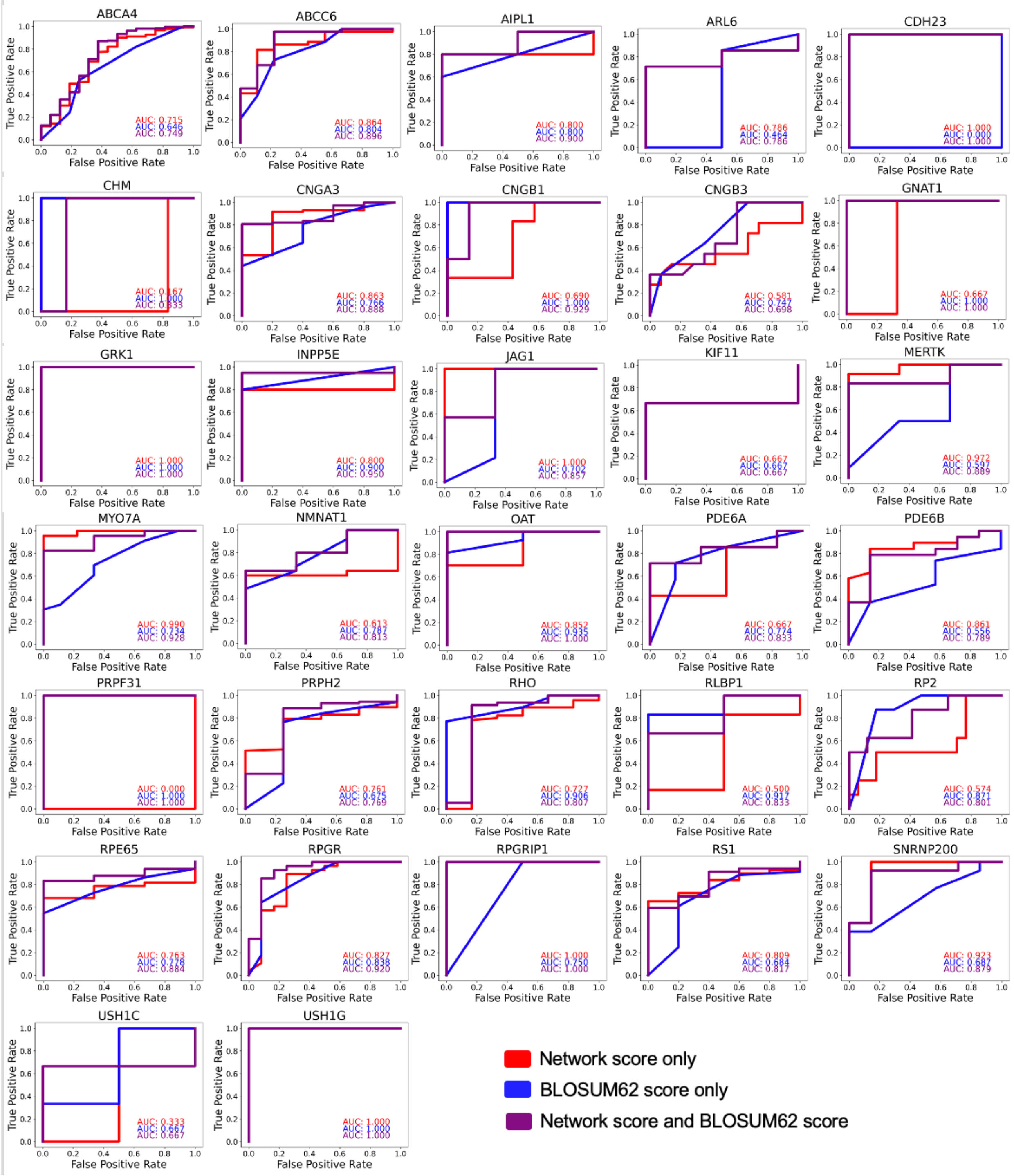


**Figure S2. Logistic regression-based modelling using SBNA and BLOSUM62 is superior to univariate models for some individual proteins.** Application of univariate and multivariable logistic regression models to the 32 inherited retinal disease proteins for which there was sufficient data to facilitate individual analysis. All regressions were trained on all proteins except the protein of interest and then tested on that protein. AUC values are shown.

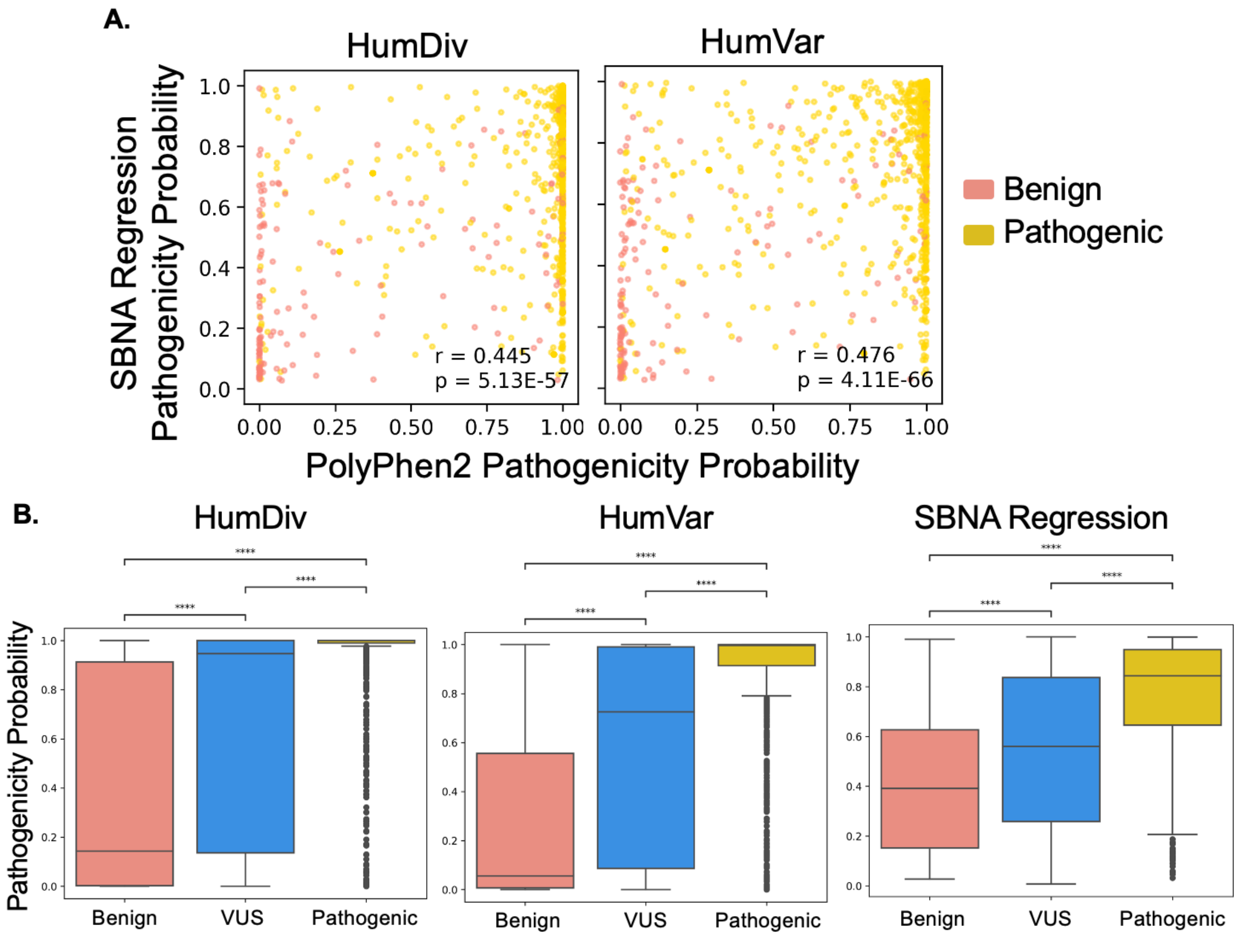


**Figure S3. SBNA regression estimates show similar trends to estimates from PolyPhen2.** **(A)** Comparison between pathogenicity probability estimates generated by the SBNA regression and PolyPhen2 trained on either the HumDiv or HumVar training data, with Spearman correlation coefficients displayed for each plot. **(B)** Comparison between pathogenicity probability estimates grouped by benign, VUS, and pathogenic variants as determined by ClinVar and gnomAD.

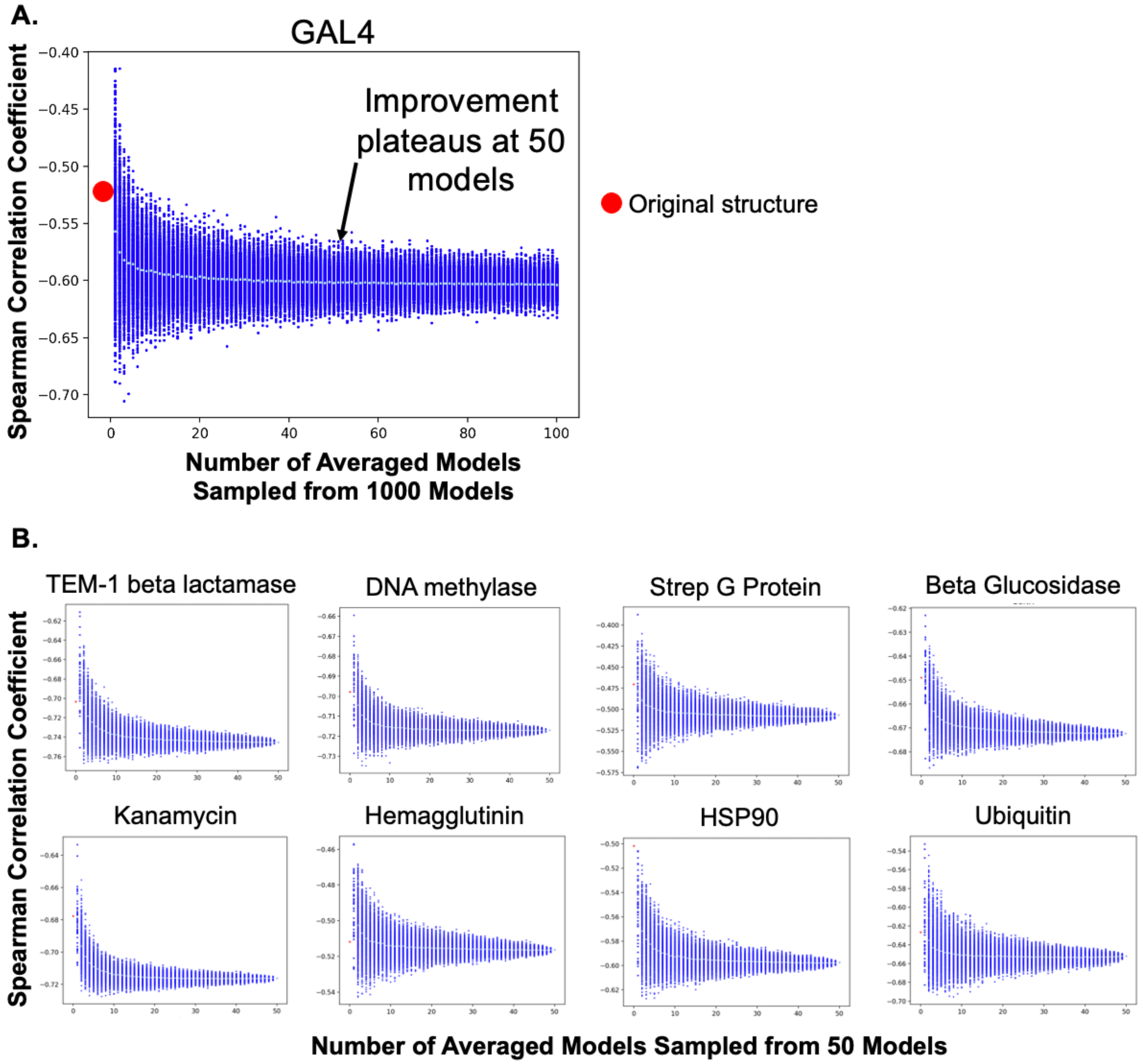


**Figure S4. Homology modelling of benchmark proteins shows improvement in correlation between network scores and functional scores. (A)** A threshold of 50 homology models was determined to be the point beyond which there was limited further improvement the Spearman correlation coefficient between network scores and functional scores from *in vitro* assays. **(B)** The effect of homology modelling on the Spearman correlation coefficient between network scores and functional scores from *in vitro* assays was measured across 8 additional benchmark proteins. Red dots depict the Spearman correlation coefficient for the original structure without homology modelling.

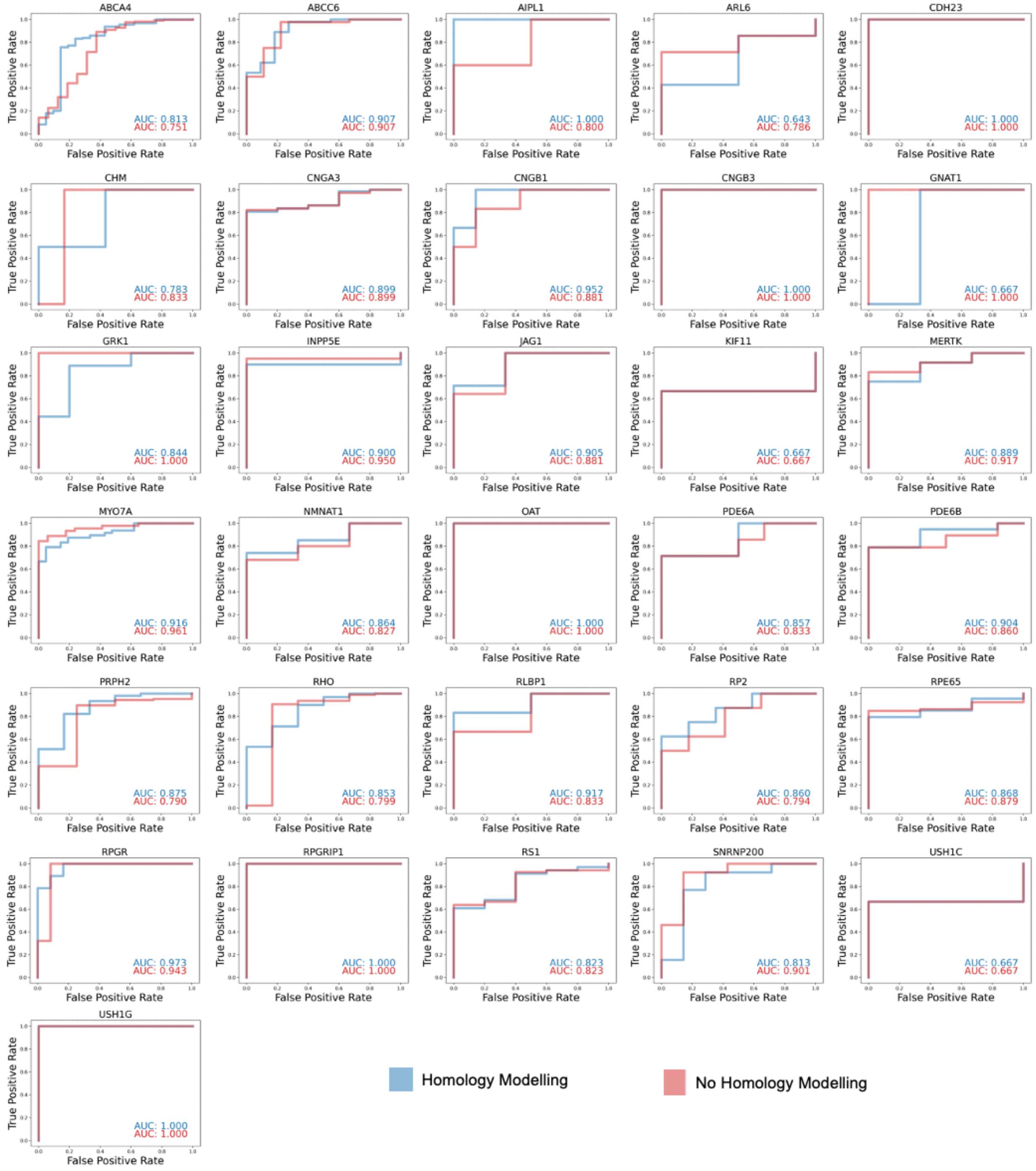


**Figure S5. Multivariable logistic regressions with and without homology modelling show similar results.** Application of the multivariable logistic regression model incorporating network scores and BLOSUM62 scores to 31 inherited retinal disease proteins. All regressions were trained on all proteins except the protein of interest and then tested on that protein. AUC values are shown.

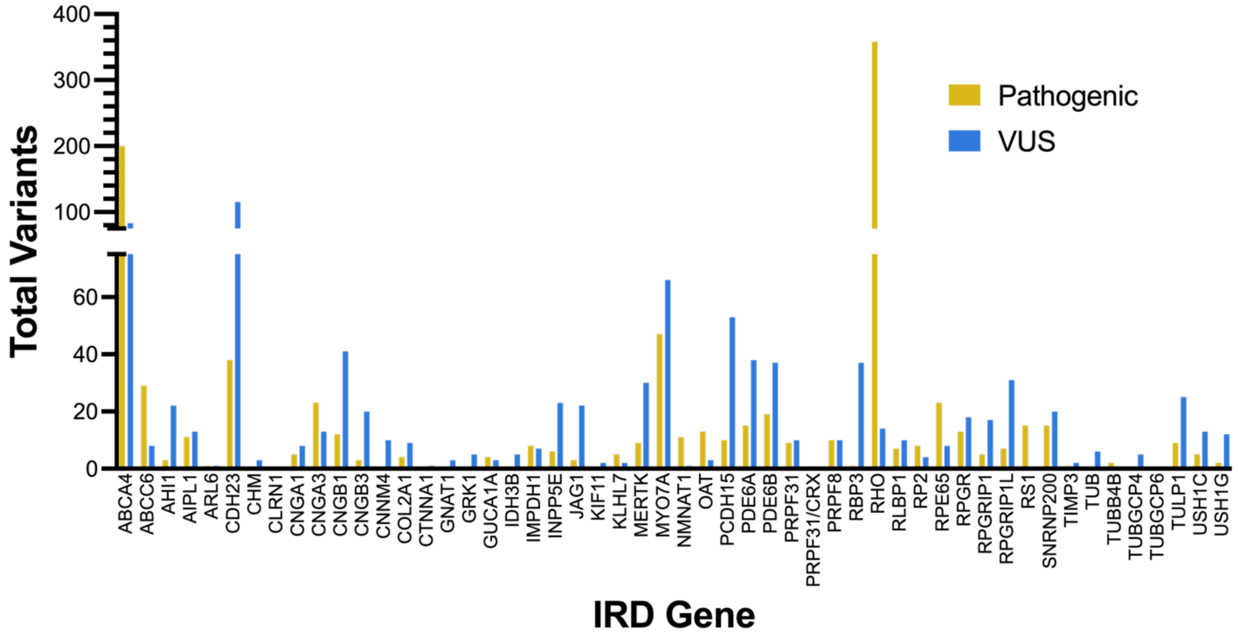


**Figure S6. Pathogenic variants and VUS from MEE patients span 52 IRD genes.**

Distribution of total pathogenic variants and VUS (as categorized by ClinVar) from MEE patients across the 52 IRD genes considered in this analysis.

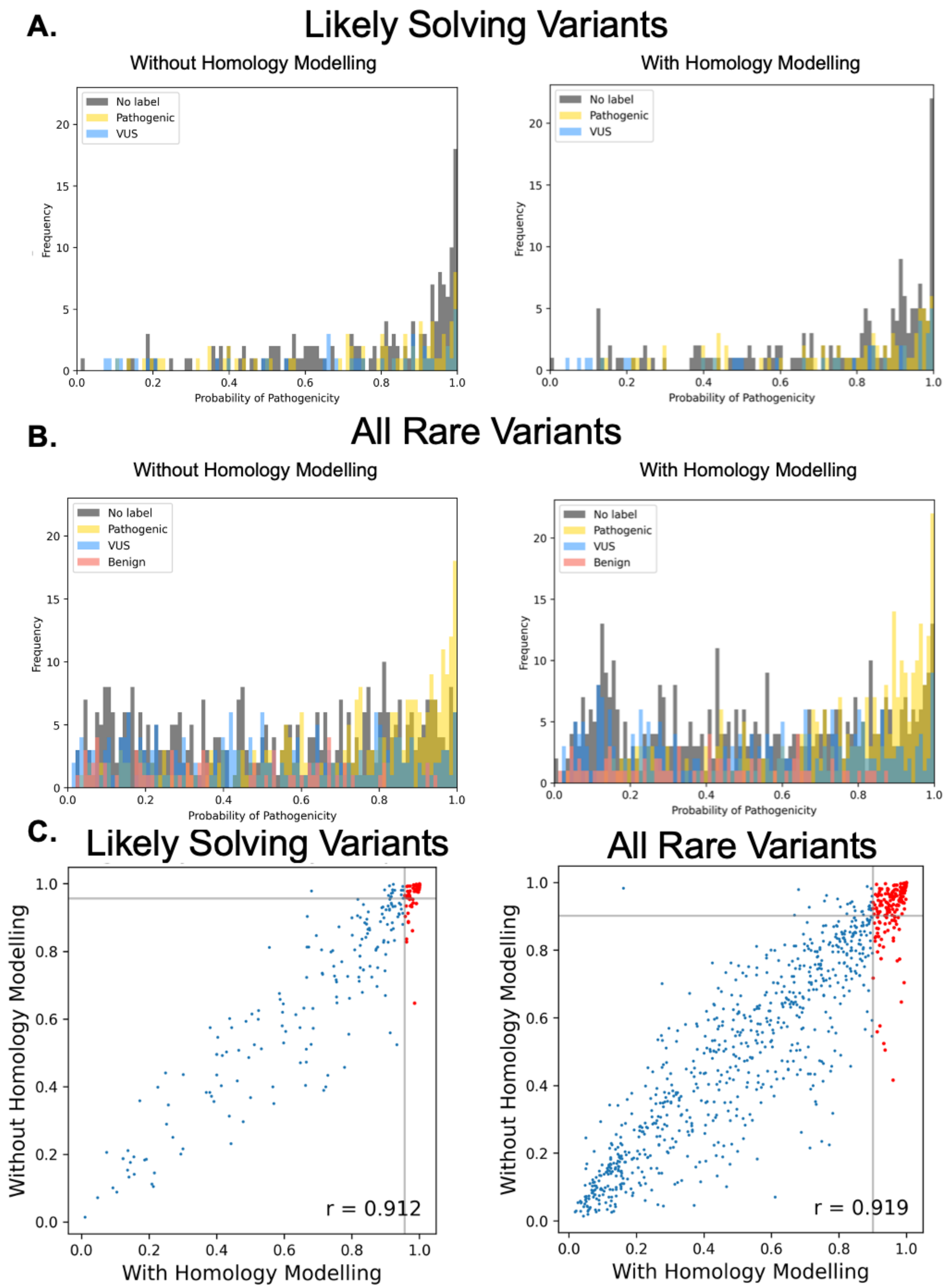
A.
Likely Solving Variants
Without Homology Modelling
With Homology Modelling
No label
Pathogenic
VUS
Frequency
Probability of Pathogenicity
B.
All Rare Variants
Without Homology Modelling
With Homology Modelling
No label
Pathogenic
VUS
Benign
Frequency
Probability of Pathogenicity
C.
Likely Solving Variants
All Rare Variants
Without Homology Modelling
With Homology Modelling
r = 0.912
r = 0.919

**Figure S7. Unsolved patient variant pathogenicity probabilities are similar with and without homology modelling.** Pathogenicity probability distributions by variant label for the **(A)** "likely solving variants" and **(B)** "all rare variants" datasets. **(C)** Pathogenicity probability comparisons with and without homology modelling. Red dots indicate variants that were included in the list of top hits for further analysis. Spearman correlation coefficients are show and are statistically significant with $p < 0.0001$.

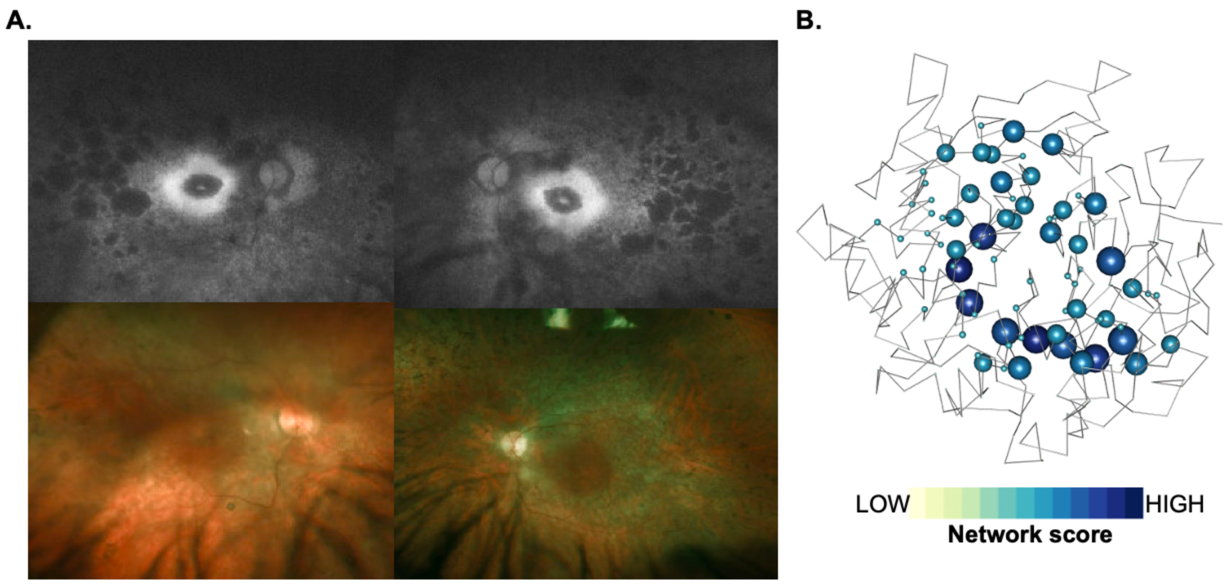


**Figure S8. SBNA helps identify pathogenic variants in a patient with RPGR-related inherited retinal disease.** Representation of network scores for a sample structure with putative solving genetic mutations. Sphere radius corresponds to network score magnitude at a particular position. A patient with clinical evidence of RPGR-related disease **(A)** but with no complete genetic explanation was fully solved using SBNA which highlighted a hemizygous mutation (Cys302Tyr) that score highly in the RPGR protein structure **(B)**.

| Protein | PDB |
|---|---|
| AHI1 | 4ESR |
| ARL6 | 2H57 |
| CNGA1 | 7LFT |
| CNNM4 | 6G52 |
| CNNM4 | 6RS2 |
| COL2A1 | 5NIR |
| GRK1 | 3C4Z |
| GUCA1A | 2R2I |
| IMPDH1 | 7RER |
| INPP5E | 2XSW |
| JAG1 | 4CC0 |
| KIF11 | 1Q0B |
| KLHL7 | 3II7 |
| MERTK | 7AB0 |
| NMNAT1 | 1KKU |
| OAT | 1OAT |
| OFD1 | 6E0T |
| PRPF8 | 3ENB |
| RLBP1 | 3HY5 |
| RP2 | 2BX6 |
| RS1 | 3JD6 |
| SNRNP200 | 4KIT |
| TUB | 1S31 |
| TULP1 | 3C5N |
| PTEN | 1D5R |
| HRAS | 4NIF |
| ABCA4 | 7LKZ |
| ABCC6 | 6BZS |
| ABCC6 | 6BZR |
| AIPL1 | 6PX0 |
| CDH23 | 5TFM |
| CDH23 | 5WJ8 |
| CDH23 | 5VVM |
| RPGR | 4QAM |
| RPGRIP1 | 4QAM |
| CTNNA1 | 4IGG |
| CHM | 1VG9 |
| CHM | 1VG0 |

| | |
|---|---|
| CNGA3 | 7RHS |
| CNGB1 | 7RH9 |
| CNGB3 | 7RHS |
| DFNB31 | 6KZ1 |
| DFNB31 | 6FDD |
| DFNB31 | 6FDE |
| ERCC6 | 7OO3 |
| GNAT1 | 1TND |
| GNAT1 | 1TAD |
| GNAT1 | 1TAG |
| IDH3B | 6KDF |
| MYO7A | 5MV9 |
| MYO7A | 3PVL |
| PCDH15 | 5ULY |
| PCDH15 | 6E8F |
| PCDH15 | 5T4M |
| PCDH15 | 4XHZ |
| PDE6A | 6MZB |
| PDE6B | 6MZB |
| PRPF3 | 6QW6 |
| PRPF31 | 2OZB |
| RBP3 | 1J7X |
| RBP3 | 4LUR |
| TIMP3 | 3CKI |
| USH1C | 3K1R |
| USH1G | 3K1R |
| BRCA1 | 1JM7 |
| BRCA1 | 1T29 |
| ERK2 | 4FMQ |
| ERK2 | 4NIF |
| RPE65 | 3FSN |
| RPE65 | 3KVC |
| RPE65 | 4F2Z |
| RHO | 1GZM |
| RHO | 3CAP |
| PRPH2 | 7ZW1 |

**Table S1. Protein Data Bank accession numbers for well-studied human proteins and IRD genes.** Protein Data Bank[31] accession numbers listed here were used to access structural data for well-studied human proteins and IRD proteins.
